# Usability and workflow implications of an electronic health record-integrated tool for automated phenotyping: Formative evaluation of Pheno+

**DOI:** 10.64898/2026.09.22.26363702

**Authors:** Peter Taber, L Weaver, Tony Di Sera, Chelsea Solorzano, Emerson Lebleu, Mickey Bolyard, Jorie Butler, Tanner Ellsworth, Isabelle Cooperstein, Paul Estabrooks, Kensaku Kawamoto, Phillip B. Warner, Kelsey Simek, Martin Tristani-Firouzi, Alistair Ward, Douglas Martin, Sabrina Malone Jenkins

**Author notes:** Corresponding author: Peter Taber.

## Abstract

**Objective:** To evaluate usability and workflow implications of Pheno+, an electronic health record- (EHR) embedded natural language processing tool that extracts patient phenotypes and maps them to Human Phenotype Ontology (HPO) terms.

**Materials and Methods:** This study performed a mixed-methods formative evaluation of Pheno+ in the context of rapid genome sequencing in the neonatal intensive care unit. Two rounds of think-aloud interviews and surveys were conducted with neonatologists, advanced practice providers, medical geneticists, genetic counselors, non-genetics pediatric subspecialists and laboratory molecular geneticists. Usability was assessed using the System Usability Scale (SUS). Structured data were analyzed descriptively. Qualitative data underwent hybrid deductive-inductive thematic analysis.

**Results:** Twenty-eight interviewees participated in two rounds of data collection. Users retained an average of 24.2 HPO terms from an average of 45.6 terms per patient. Mean time curating terms was 8:59 per patient. Final SUS score was 76 (above scale mean). Qualitative themes included: burden of curating terms; interpretation of phenotypes; adding information to phenotypes; role of genetics expertise; and tool uses beyond the original core use case for Pheno+.

**Discussion:** Pheno+ demonstrated positive usability but required curation effort to reduce erroneous terms. Participant responses indicate that effective use may depend on knowledge of HPO structure and laboratory workflows. Automated phenotyping seems likely to reduce net clinician workload, while also reshaping clinical responsibilities, expertise requirements, and communication in genetic testing.

**Conclusion:** Tools like Pheno+ can support scalable genomic medicine. Design and implementation may benefit from addressing curation burden, phenotype contextualization, expertise requirements, and other workflow needs.

## Introduction

The construction of a patient’s phenotype for genetic testing has traditionally involved manual review of the patient’s chart, requiring substantial clinician time and expertise. At the same time, accurate phenotyping is critical for genetic test selection and accurate diagnosis, making phenotyping a major bottleneck in genetic testing workflows.^1,2^ Automated tools for phenotyping offer an important solution for scaling genetic testing in routine clinical practice. These tools typically automate two important steps of the phenotyping workflow: first, they segment and extract phenotype information from the patient’s chart; second, they map this information to items in controlled vocabularies such as the Human Phenotype Ontology (HPO).^3–5^ The resulting standardized phenotypes may guide test selection (e.g. gene panel selection or whole-genome sequencing)^6^, support algorithm-based variant prioritization^7,8^, and guide interpretation of results.^9^

A growing literature demonstrates the feasibility of using a range of rule- and large language model- (LLM-) based natural language processing (NLP) tools in automated phenotyping, often showing significant time-savings as well as noninferior or superior accuracy compared to manual chart review.^10^ These gains are highly promising and urgently needed to address the challenge of scaling genomic medicine to meet growing demand.^11^ However, deployment of these tools and implementation in real-world, clinical workflows requires consideration of substantial additional factors. Research in human factors engineering and sociotechnical systems has revealed that quantitative gains in efficiency afforded by automation may be accompanied by qualitative changes in overall system functioning.^12,13^ For example, automation may require clinicians to perform new tasks (e.g. monitoring automation performance) and/or gain novel expertise (e.g. knowledge of how to interpret machine output or maintain automated systems). Research on other complex work systems suggests that envisioning and planning for changes induced by automation in genetic testing pathways may be crucial for managing the transition to scalable, artificial intelligence-supported genomic medicine.^14–16^ Studies of workflow and implementation of automated phenotyping tools to support such planning remain limited.

With this gap in mind, this formative evaluation studied the automated phenotyping tool Pheno+. Pheno+ is an NLP-based tool using HPO terms, piloted in the context of rapid genome sequencing (rGS) in the University of Utah neonatal intensive care unit (NICU). Here, we focus on the tool’s usability, broader fit with existing clinical workflows and interviewees’ visions of its potential future use, with the ultimate goals of supporting tool adoption and ensuring high diagnostic yield in future clinical genetic testing. Quantitative analyses of tool-supported phenotyping efficiency and quality compared to manual phenotyping will be reported separately. The evaluation is part of a larger parent study seeking to implement novel informatics tools into NICU rGS workflows.

## Materials and Methods

### Overview

We conducted a mixed methods formative evaluation of the novel clinical phenotyping tool Pheno+. Data collection methods included two rounds of think-aloud interviews focused on phenotyping tasks using the tool, and two corresponding rounds of post-interview surveys. In accordance with the Common Rule, the research was submitted to the University of Utah’s Institutional Review Board and exempted (IRB_00165986).

### Pheno+ Structure

Pheno+ is an electronic health record- (EHR-) integrated clinical phenotyping tool that extracts phenotype information from patient notes and maps it to HPO terms for clinician review. It is implemented using the Substitutable Medical Applications and Reusable Technologies (SMART) framework within Health Level 7 Fast Health Interoperability Resources (FHIR)^17^, a vendor-neutral standard for launching applications from within the EHR, authenticating users via Open Authorization (OAuth) 2.0, and accessing patient data through FHIR Release 4^18^ interfaces. Pheno+ launches directly from the patient chart in the Epic EHR platform and receives scoped, read-only access to the active patient’s clinical notes.

Pheno+ is comprised of a client application and a phenotype extraction service. The client is a single-page web application (Vue.js) that runs within the EHR, retrieves clinical notes as FHIR DocumentReference resources^19^, and presents the review interface. Phenotype extraction is performed by a stateless server-side service, hosted within the institution’s protected environment, that accepts note text and returns extracted HPO term identifiers along with the source text block supporting each term. At the time of evaluation, the extraction service used ClinPhen^20^, a rule-based NLP tool that allows for deployment on commonly available healthcare infrastructure. ClinPhen matches note text against a lexicon of HPO term names and synonyms chosen for its light hardware requirements, and operates independently of the client application, allowing ClinPhen to be replaced by other phenotyping tools^21–24^ without modifying the EHR integration or user interface. HPO term identifiers are resolved to names in the browser using a bundled copy of the HPO, which also supports type-ahead search for terms not detected by the extraction engine. Licensing, availability and technical information about Pheno+ can be found at its Github repository (https://github.com/iobio/pheno-plus). The user interface can be seen in Figure 1.

**Figure 1.**
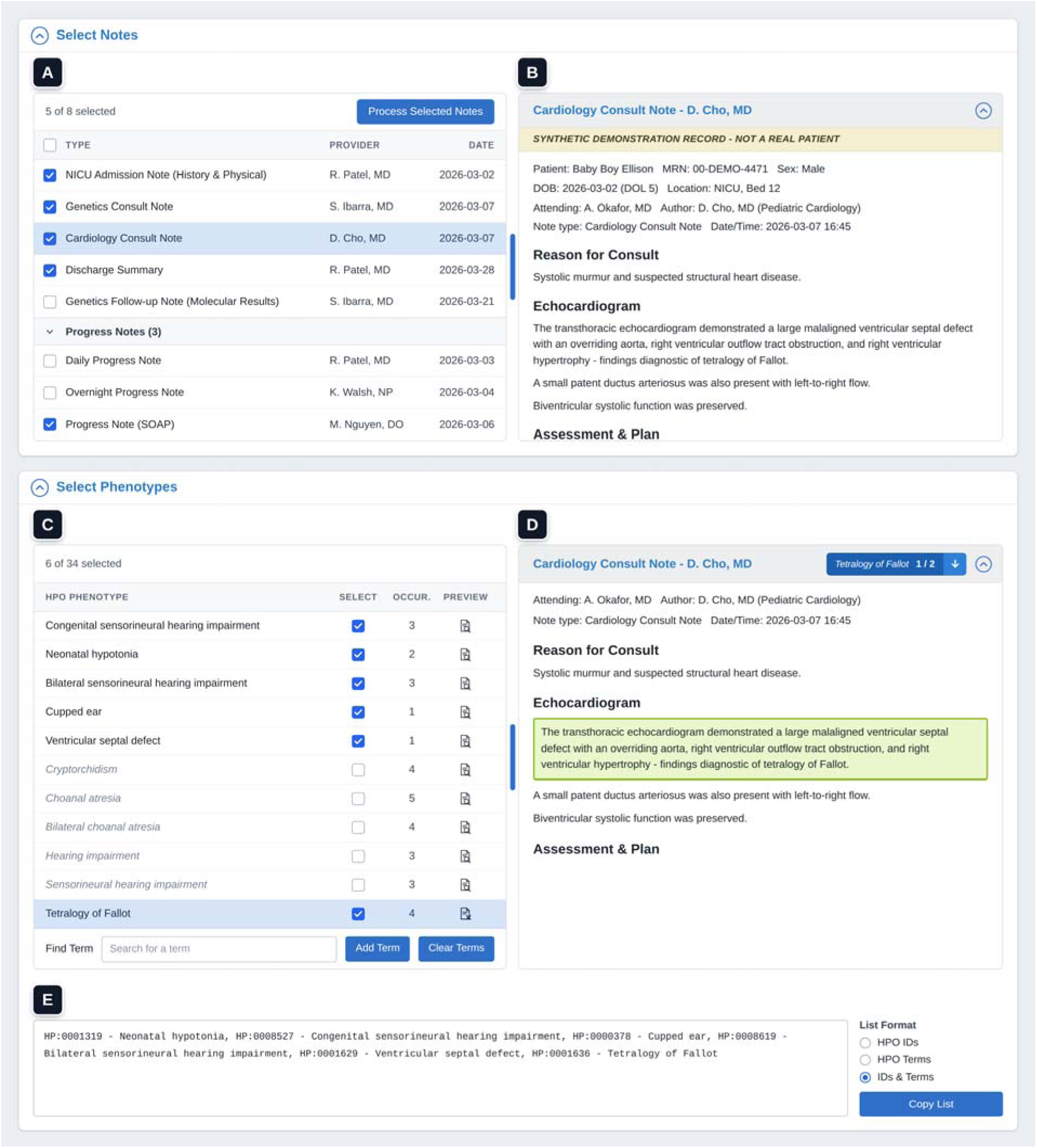
User interface of Pheno+, including available notes for process (section A), the content of the selected note (section B), terms generated after processing notes (section C), highlighted relevant portion of a note supporting a term generated by the tool after processing (section D), and the list of HPO terms with unique identifiers (section E).

### Pheno+ Workflow

A phenotyping session has four steps. First, after launch, Pheno+ displays the patient’s clinical notes, restricted to note types most likely to describe the phenotype, such as admission histories and physicals, consultation notes, progress notes, and discharge summaries (Figure 1A). Opening a note displays its text alongside the list (Figure 1B). The user then selects notes to analyze. Second, Pheno+ sends the text of the selected notes to the extraction service, which returns a list of HPO terms, each with its name and a count of supporting mentions (Figure 1C). This automates the first pass of phenotyping, which otherwise requires reading each note and coding findings to HPO by hand. Third, the user curates the list, determining which terms best describe the patient’s phenotype and searching the ontology to add any that extraction missed. Where a term warrants closer review, the user can open it to highlight its supporting mentions in the source note and step through them, checking the extraction against the original documentation (Figure 1D). Finally, the user copies the completed list to the clipboard (Figure 1E). This curated list of HPO terms accompanies the sequencing requisition for the reference laboratory to incorporate into its analysis, and serves as structured input to pre- and post-sequencing computational tools.

### Data Collection

Data collection occurred in two rounds. Across both rounds, roles recruited included neonatologists and advance practice providers (APPs), medical geneticists, non-genetics subspecialists, genetic counselors, and laboratory molecular geneticists. Because they play a similar role as the responsible attendings for NICU patients, neonatologists and APPs were treated as a single role in analysis. In both rounds, participants were prompted to follow the general workflow described above.

In Round 1, interviewees were grouped by role and performed a think-aloud phenotyping task together on a single real patient in the Epic electronic health record (EHR) platform. This interview round was intended to provide rapid feedback to guide coarse changes in the tool’s appearance and functioning, which were then implemented by the design team. In Round 2, individual interviewees completed three phenotyping tasks on patients drawn from the bottom, middle and upper tercile of patient complexity, using the number of available notes as a proximate measure of complexity (n=58 patients). Within terciles, patients were assigned to interviewees randomly. In both rounds, interviewees completed a post-interview survey about the tool, including the System Usability Scale (SUS)^25^, as well as open-ended responses to explain their ratings. Interviews were performed using the Zoom platform and digitally recorded. Survey data were collected via the Qualtrics platform.

### Data Analysis

Structured data were extracted from video recordings of Round 2 interviews. Video data and SUS survey data were summarized in Excel.

Interviews from both Rounds were professionally transcribed. Transcripts were thematically analyzed using the qualitative analysis platform Dedoose (SocioCultural Research Consultants, Redondo Beach).^26^ Thematic analysis was initiated by familiarizing the full team with the corpus, followed by memoing and coding. A hybrid deductive-inductive approach was used to code transcripts. The codebook was initiated using a priori concepts relevant to formative evaluation (e.g. perceived reduction in task burden, feature requests), as well as constructs derived from the integrated Promoting Action on Research Implementation in Health Sciences (i-PARIHS) framework guiding the parent project (e.g. codes related to intervention and recipient characteristics).^27^ Additional codes were added based on initial memoing and emergent insights from the corpus (e.g. suggestions about unforeseen ways to use Pheno+ in existing workflows; variations in interpretations of the tool output). Codes were then considered as candidates to be aggregated in themes based on topical relevance and ability to provide insight into the usability and workflow issues of central concern to this study.

## Results

Here, we describe our results in terms of 1) participant recruitment; 2) tool revisions; 3) tool performance and user interactions; and 4) qualitative feedback on tool use and workflow implications.

*1) Participant recruitment:* A total of eight individuals participated in Round 1 group interviews and surveys. Twenty individuals participated in Round 2 interviews and surveys. Table 1 describes interviewees’ clinical roles and years of experience for both Rounds.
*2) Tool revisions:* Round 1 yielded feedback to improve querying clinical notes and management of the tool’s views, such as the ability to collapse and resize windows. Pheno+ was also revised to display the approximate number of “mentions” supporting each HPO term in its output in order to improve the transparency of the tool’s recommendations. Additional insights on the Pheno+ design process will be reported in a companion piece.
*3) Tool performance and user interactions:* The average number of notes per patient for all patients was 7.6. Users retained an average of 76.5% of a patient’s notes for processing by Pheno+. Pheno+ generated an average of 45.6 HPO terms per patient case, with users retaining an average of 53% of those terms in their final phenotypes. Total time spent reading and selecting notes to include averaged 1:41 per patient for all roles. Total time spent examining and selecting terms to retain in the final phenotype averaged 8:59 for all roles.

**Table 1.** Participant characteristics for data collection in Round 1 (group interviews) and Round 2 (individual interviews). APP=advanced practice provider; St.Dev.= standard deviation.

| Round | Role | N | Mean Years in Role (St. Dev.) |
| --- | --- | --- | --- |
| 1 (n=8) | Neonatologist/APP | 2 | 19 (8.5) |
|  | Non-genetics subspecialist (pediatric gastroenterologist and neurologist) | 2 | 7.5 (3.5) |
|  | Genetic counselor | 2 | 1.5 (0.7) |
|  | Medical geneticist | 2 | 10 .0 (11.3) |
| 2 (n=20) | Neonatologist/APP (including two Fellows) | 8 | 9.1 (7.7) |
|  | Non-genetics subspecialist (two pediatric neurologists) | 2 | 9.0 (8.5) |
|  | Genetic counselor | 3 | 7.3 (5.9) |
|  | Medical geneticist (including two Fellows) | 4 | 9.5 (8.2) |
|  | Molecular geneticist (including two Fellows) | 3 | 10.3 (12.9) |

As genetic testing increasingly extends beyond genetics specialists into broader clinical practice, differences in performance between clinicians with and without specialized genetics training are particularly relevant. Neonatologists and APPs spent significantly less time curating terms than clinicians with specialized genetics training (mean=6:42 versus 10:31, p=.029). However, when all clinicians without specialized genetics training were considered together, curation time did not differ significantly from that of genetics specialists.

Finally, for usability calculations, eight responses were recorded in Round 1. One Round 2 survey response was excluded due to incomplete data, leaving a total of 19 Round 2 respondents. Surveys in Rounds 1 and 2 yielded SUS scores of 78 and 76 (both good to excellent), respectively. Table 2 provides an overview of structured data for all roles in Round 2.

*4) Qualitative feedback on tool use and workflow implications:* Via thematic analysis of Round 1 and Round 2 interviews, as well as unstructured survey responses, we identified five key themes of users’ feedback and commentary about the tool, discussed below.

**Table 2.**
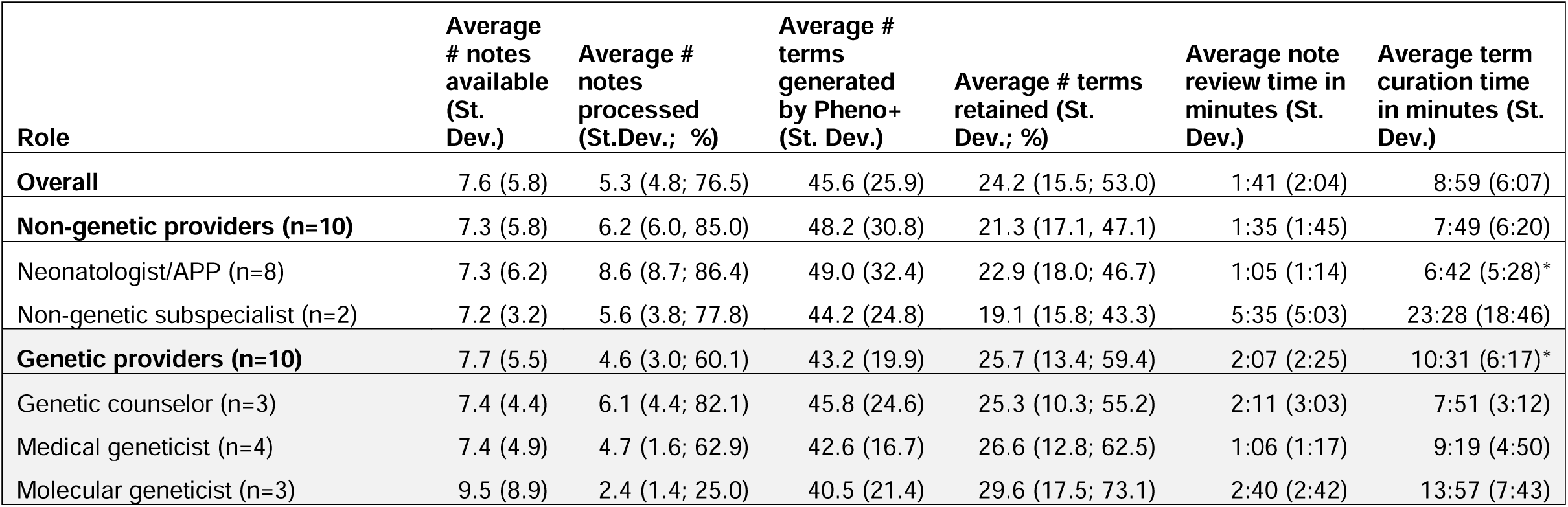
Structured data for all Round 2 phenotyping tasks. APP=advanced practice provider; St.Dev.=standard deviation; *=statistically significant difference (p=.029)

### I) Clinicians emphasized the burden of identifying and removing erroneous HPO terms

While overall attitudes toward Pheno+ tended to be positive, participants routinely noted dissatisfaction with the curation effort required to identify and remove terms in the output list that they deemed erroneous. As noted above, the majority of participants’ time during the phenotyping task was spent curating output from Pheno+, which involved examining which terms the tool had generated and reading some portion of the term’s supporting text in the clinical notes, with the goal of confirming the validity of the term. Discerning the clinical relevance of a term is a complex task, as demonstrated in Excerpt 1. The high volume of terms that were identified as erroneous contributed to reduced appraisals of usability, as in Excerpt 2. Terms deemed erroneous often derived from the provider’s differential (Excerpt 3), or from conditions that clinicians wanted to monitor the patient for but did not confirm at the time of writing the note (Excerpt 4). Erroneous terms could also derive from the maternal or family histories, from descriptions of care processes like rounding, the administration of vaccines, or other note content.

### II) Clinicians used disease scripts and clinical context to assess diagnostic relevance of HPO terms

Clinicians often reasoned through HPO terms’ potential relevance to an ultimate rGS diagnostic outcome based on the typical course and symptomology of diseases they had familiarity with. For example, in Excerpt 5 the provider asserts the importance of generalized hypotonia to the patient’s condition after examining their notes and then speculates about the potential relevance of cephalohematoma to the phenotype, offering Noonan’s syndrome as one potential diagnosis for which this term would be relevant.

Terms with potential genetic relevance required interpretation to evaluate whether they referenced a symptom of a primary condition of interest. For example, in Excerpt 6 the neonatologist describes identifying a primary condition with unclear genetic relevance and then including or excluding additional HPO terms based on their relationship to the primary condition. Clinicians varied in their approaches to the inclusion of HPO terms for secondary symptoms in their final phenotypes. While terms perceived to be secondary were often excluded (Excerpts 7, 8, and 9), they might also be included, depending on whether the clinician believed it advantageous to take a more exhaustive approach. For example, in Excerpt 10 a clinician sees the term “hyperbilirubinemia”, questions whether the term is relevant, searches the baby’s chart and finds that treatment was administered for the condition, then decides to include the term while expressing doubt that the hyperbilirubinemia is directly related to any underlying genetic disorder. Finally, clinicians also judged terms’ relevance in the broader context of the NICU, for example when considering that uncertainty over terms’ relevance might resolve over the course of a neonate’s development (Excerpt 11).

### III) Clinicians expressed interest in enriching the HPO-based phenotype with additional information

Clinicians often suggested that adding context or interpretive clues to the HPO term list would be valuable. The simplest request, made by several clinicians, was the ability to flag specific terms as high priority, as in Excerpt 12. A medical geneticist noted that providing both an exhaustive list and a separate list of prioritized terms was their current standard practice (Excerpt 13). Some clinicians desired the ability to indicate uncertainty about terms’ relevance (Excerpt 14). One clinician noted that, in certain cases, terms deriving from the mother’s or family history might also be valuable to include (e.g. for families opting in to secondary findings; Excerpt 15). Additionally, some clinicians indicated a desire to capture HPO terms or information from structured documentation, imaging or lab reports not parsed by the tool at the time of evaluation, along with the HPO-based phenotype (Excerpt 16). One molecular geneticist emphasized that difficulties communicating prioritized HPO terms between hospital and laboratory was a characteristic problem of using the HPO (Excerpt 17).

### IV) Specialized knowledge of genomics, test procedures and the HPO played an important role in expert users’ strategies to curate tool output

Pheno+ users with more extensive knowledge and experience using HPO and genetic testing were able to use that knowledge to shape their strategies for term inclusion. In Excerpt 18, in the context of debating what the patient’s primary condition is, a pediatric neurologist notes that inclusion of congenital myopathy could have important implications for genetic testing (e.g. for test selection). The determination of the appropriate level of generality of HPO terms was another issue raised. In Excerpt 19, a genetic counselor considered the general tension between balancing breadth (to ensure that pathogenic variants are not missed by the laboratory) and specificity (to avoid excessive search effort for laboratory geneticists) while curating terms. Appropriately selecting terms at the right level of the ontology could require consideration of condition-specific factors. For example, in Excerpt 20 a medical geneticist shared the insight that including only highly specific terms to describe congenital heart defects could easily lead to missed variants. The complexities of term selection led some clinicians with deeper background in genomics to emphasize the need for non-specialist users to have expert oversight, as in Excerpt 21.

### V) Suggested use cases for the tool extended beyond pre-test phenotyping

Genetic counselors noted that the tool could help them respond to frequent requests by the care team after test send-out for reminders of the exact phenotype that was provided to the laboratory (Excerpt 22). Beyond the immediate setting of the NICU, the tool was also seen as facilitating synopses of new patients with longer medical histories (Excerpt 23) and overviews of phenotype changes for patients beyond their hospitalization (e.g. in the context of decisions about reanalysis; Excerpts 24 and 25). Having the phenotype viewable within clinical documentation was seen as a potentially valuable step to make the rGS process more transparent (Excerpt 26). One genetic counselor suggested that curating terms to create a persistent phenotype as a part of clinical documentation in the EHR would ideally involve greater selectivity for HPO terms compared to constructing the phenotype for the sole purpose of aiding laboratory analysis (Excerpt 27).

## Discussion

This study conducted a formative evaluation of Pheno+, an EHR-embedded, NLP-based phenotyping tool using HPO terms to create standardized phenotypes. The study focused on the use case of past NICU patients for whom rGS had been sent at the time of tool evaluation, providing insights into usability as well as the dynamics of workflow integration for Pheno+ or similar tools in regular clinical practice.

We found that user experience was impacted by the need to remove terms that were deemed erroneous or irrelevant to a genetic workup for the patient at hand. We note that at the time of evaluation Pheno+ relied on rule-based NLP. Multiple approaches have been developed for automated HPO term extraction, spanning dictionary-based methods^23,28^, transformer-based concept recognition^24^, and retrieval-augmented LLM approaches^21,22^. Future work will explore the integration of more advanced models to improve performance in Pheno+. Since even a high-performing LLM-based tool is unlikely to perform perfectly, transparency about the need to monitor phenotype quality, as well as co-designed workflows to ensure phenotype quality may be important for future implementation. These strategies can help shape users’ “performance expectancy” for the tool (expectations about its effectiveness, including its error prone-ness), and the “effort expectancy” of using it (expectations about how laborious it will be to use), both key factors conditioning the adoption of technology-based interventions.^29–32^

For the core use case of constructing a phenotype prior to test send-out, a well-constructed phenotype communicates a patient’s presentation in clear, standardized terms to diagnosticians in the reference laboratory, allowing the laboratory to narrow the genomic search space by prioritizing genes and variants consistent with the observed phenotype. Intentionally supporting this communicative function of the phenotype may be important for real-world practice. For example, interviewees’ suggestions of the need to signal a distinction between primary and secondary HPO terms could be supported via flagging or ranking terms, grouping terms perceived to belong together, or other simple visual mechanisms. The large number of terms generated by automated phenotyping compared to manual workflows may make organizing the tool’s output to convey this “metadata” about terms’ significance particularly important.^33^

Participant responses implied an expanded role for standardized patient phenotypes if automation makes them readily available. As envisioned by some interviewees, a longitudinal phenotype record maintained in the EHR would make a standardized representation of the patient’s presentation visible to the entire care team, serving as a shared memory of the phenotype for which testing was ordered and allowing it to be updated. Such public information is a key feature of successfully coordinated teamwork often noted in human factors engineering and research on cooperative work.^34–36^ This use case would require storing multiple tool-generated phenotypes per patient in the EHR (along with date, time and the patient information processed by the tool), integration that Pheno+ lacked at the time of evaluation. Attending closely to these and other emergent applications for phenotyping tools may be valuable to evaluate both the merits and risks of novel use cases.

Congruent with the human factors engineering literature on automation, our findings suggest that automated phenotyping may induce changes in the tasks and expertise involved in genetic testing workflows. Per findings in Theme IV, users without formal training in genetics may initially lack knowledge of the basic content and structure of the HPO, as well as the skill to construct phenotypes to optimally support downstream laboratory work. In relation to the first concern, non-specialists may gain general familiarity with HPO content and structure, for example as they search for terms returned by the tool or attempt to determine whether specific terms exist for a symptom of interest in the online HPO resource, as they did during our interview sessions. To address potential gaps in non-specialist expertise, a brief introduction to the content and structure of HPO may be beneficial for non-specialists.

However, developing expertise in selecting HPO levels and terms to support optimal diagnostic yield may be more difficult. Some research suggests that laboratory workflows are perceived by clinicians as a “black box”, from which they receive test results without a clear understanding of the laboratory’s internal workings.^37,38^ Where this is true, opportunities may be limited for non-specialists to learn about downstream uses of the phenotypes they create, and to receive feedback that would allow them to learn to discern high- and low-quality phenotypes in particular clinical scenarios.^39–41^ Moreover, the observed difference in time spent curating HPO terms by neonatologists compared to specialists may be interpreted as evidence of a less skeptical attitude toward tool output on the part of non-specialists. This interpretation is based on a limited sample but, if accurate, could mean that non-specialists will be less motivated to voluntarily seek feedback on phenotype quality. Per interviewees’ suggestions, simple workflow modifications such as review by genetic counselors of phenotypes generated by non-specialists could provide efficient quality assurance.

Given that automated phenotyping may reshape clinical workflows, future implementations may benefit from drawing on frameworks that center the mutual adaptation between technology and practice throughout design and implementation stages. These include frameworks such as the non-adoption, abandonment, scale-up, spread and sustainability (NASSS) framework in implementation science^42,43^, as well as human factors engineering approaches focused on understanding the sociotechnical consequences of automation.^12–15,44,45^

While this study offers useful insights into the dynamics of automated phenotyping tools, it has limitations. First, the study was performed in a pseudo-laboratory context. In real-world practice, clinicians would have greater familiarity with patients they are actively caring for, but would also perform phenotyping under time pressure and amidst numerous other claims on their attention. Nonetheless, we suggest that our evaluation usefully highlights aspects of tool use rarely discussed in the literature. Second, Pheno+ utilized rule-based NLP at the time of evaluation.^28^ Tools using large language models to parse clinical notes into phenotypes may exhibit different performance characteristics, with potential gains in phenotype accuracy and relevance accompanied by significantly greater computational cost.^21,23^ Third, our evaluation occurred in the context of a larger project focused on diagnostic rGS in the NICU. Evaluation of phenotyping tools in other contexts (e.g. non-acute or adult use cases) might yield different insights. Despite these limitations, this study contributes to the existing literature by observing aspects of usability of an automated phenotyping tool, and changes that may result from widespread availability of automated phenotyping in clinical practice.

## Conclusion

This formative evaluation suggests that automated phenotyping via Pheno+ or similar tools can make phenotyping feasible for a wide range of non-genetic specialists and genetic specialists. At the same time, these tools seem likely to introduce new demands for clinical judgment and quality assurance. Pheno+ demonstrated favorable usability but required effort to curate automatically generated HPO terms. Our findings also indicate the importance of supporting communication of clinically meaningful distinctions about the phenotype (e.g. primary vs. secondary conditions), and the potential need for mechanisms of specialist oversight. Beyond pre-test phenotyping, users envisioned using phenotypes to support shared clinical understanding, longitudinal documentation, and reanalysis. In addition to studying NLP-based tool performance, future evaluations and implementations of phenotyping tools may benefit from examining automated phenotyping as a sociotechnical intervention with effects that extend across clinical and laboratory workflows. Accounting for the broader workflow dynamics discussed here may help to scale genomic medicine with the efficiency benefits of automation while preserving phenotype quality and diagnostic yield.

## Supporting information

Interview guide

## Data Availability

A subset of data may be made available upon request due to privacy considerations.

## Author Contributions

PT: Data curation, formal analysis, investigation, methodology, project administration, writing – original draft, writing – review and editing

LW: Formal analysis, investigation, writing – review and editing

TD: Software, writing – original draft, writing – review and editing

CS: Formal analysis, investigation, project administration, writing – review and editing

EL: Investigation, software, writing – review and editing

MB: Formal analysis, writing – review and editing

JB: Methodology, writing – review and editing

TE: Writing – review and editing

IC: Software, writing – review and editing

PE: Conceptualization, funding acquisition, resources, supervision, writing – review and editing

KK: Resources, software, supervision, writing – review and editing

PW: Software, writing – review and editing

KS: Investigation, writing – review and editing

MTF: Conceptualization, funding acquisition, project administration, resources, supervision, writing – original draft, writing – review and editing

AW: Resources, software, supervision, writing – review and editing DM: Software, writing – review and editing

SMJ: Conceptualization, investigation, supervision, writing – review and editing

## Acknowledgments

KK reports honoraria, consulting, sponsored research, licensing, or co-development in the past three years outside the scope of the submitted work, but in the general related areas of clinical informatics, standards-based interoperability, and/or artificial intelligence. These relationships are or were with Pfizer, RTI International, NORC at the University of Chicago, the University of Pennsylvania, Yale University, Custom Clinical Decision Support, the Office of the National Coordinator for Health IT (via Security Risk Solutions), MD Aware, the University of Colorado. the University of Michigan, Beckman Coulter, and Surescripts. KK is also an unpaid co-chair of the Health Level Seven International Clinical Decision Support Work Group and has helped develop a number of health IT tools which may be commercialized to enable wider impact. None of these relationships have direct relevance to the manuscript but are reported in the interest of full disclosure.

## Funding

This work was supported by funding from the National Institutes of Health (NIH) National Center for Advancing Translational Science (RC2TR004391; PIs Tristani-Firouzi and Estabrooks). A portion of SMJ’s time was supported by the NIH National Human Genome Research Institute (K08HG013111; PI Malone Jenkins). The content is solely the responsibility of the authors and does not necessarily represent the official views of the NIH.

---

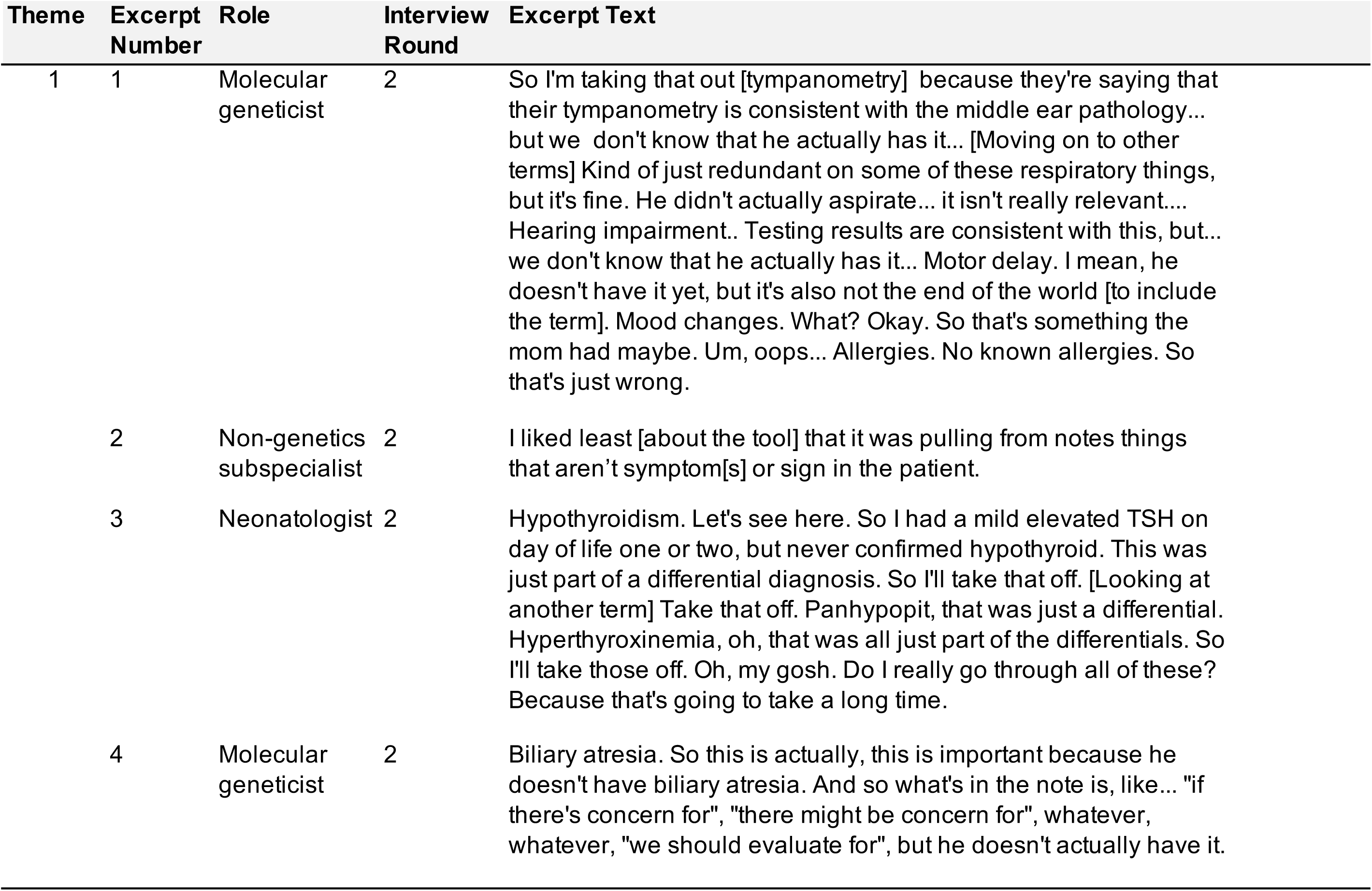
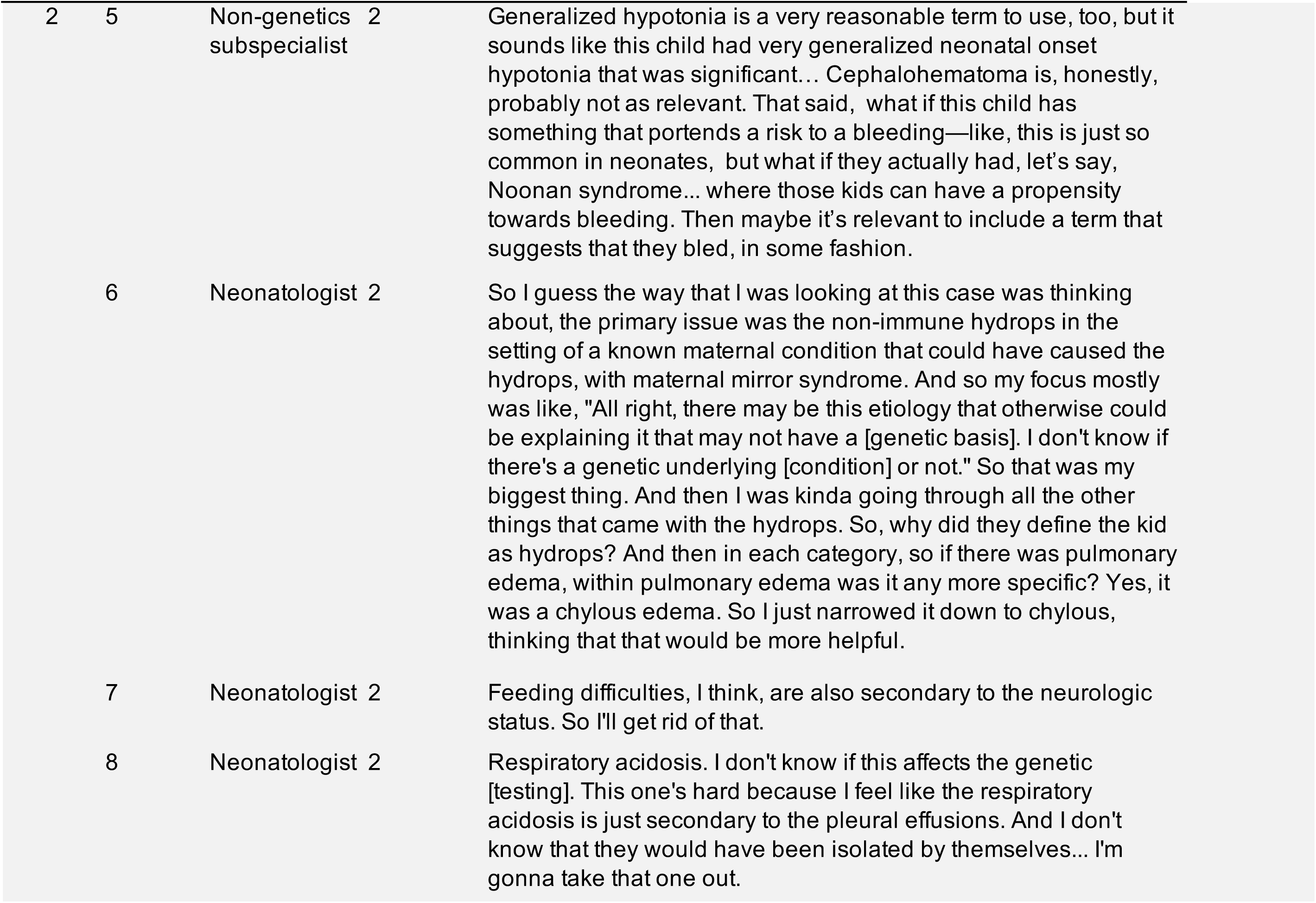
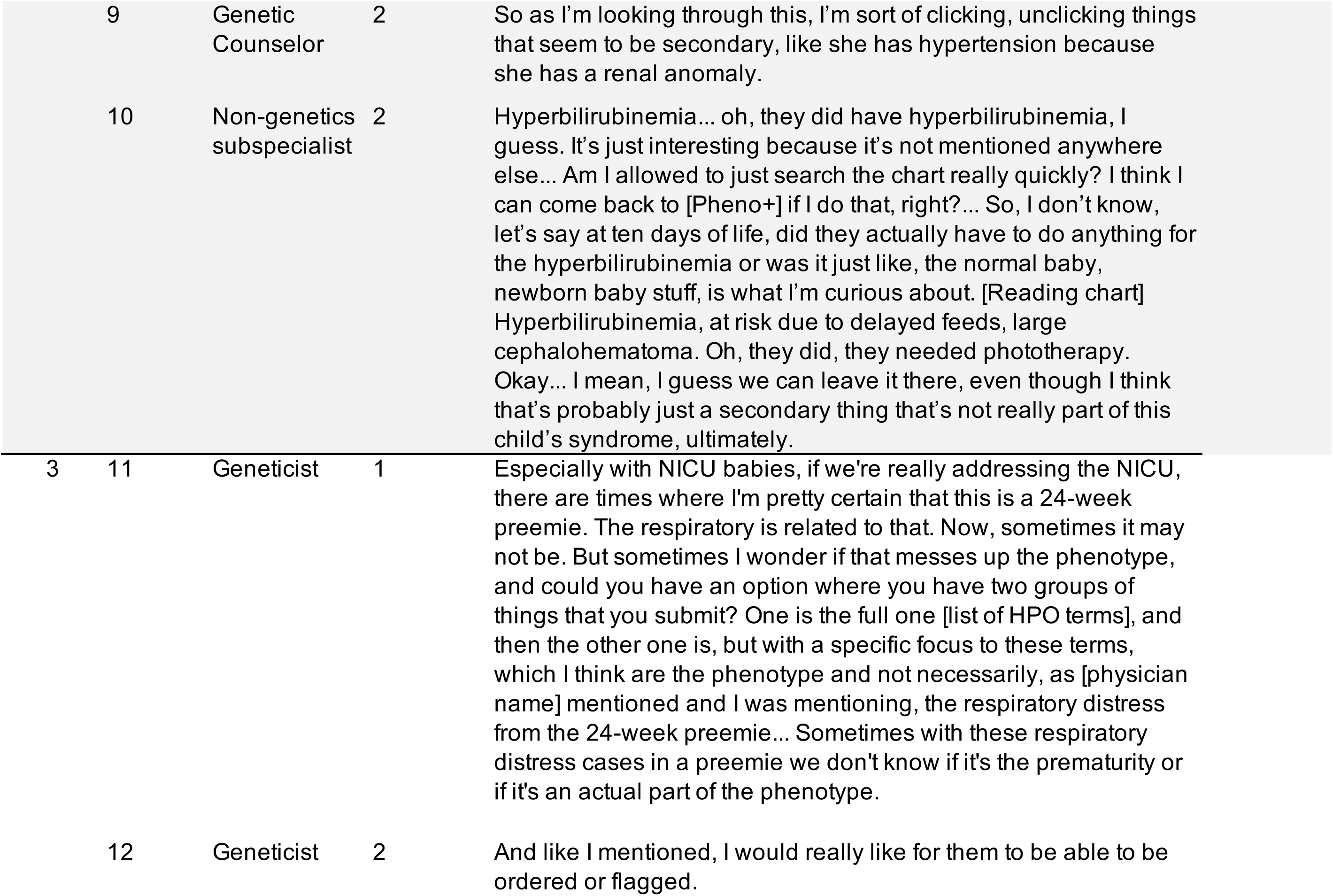
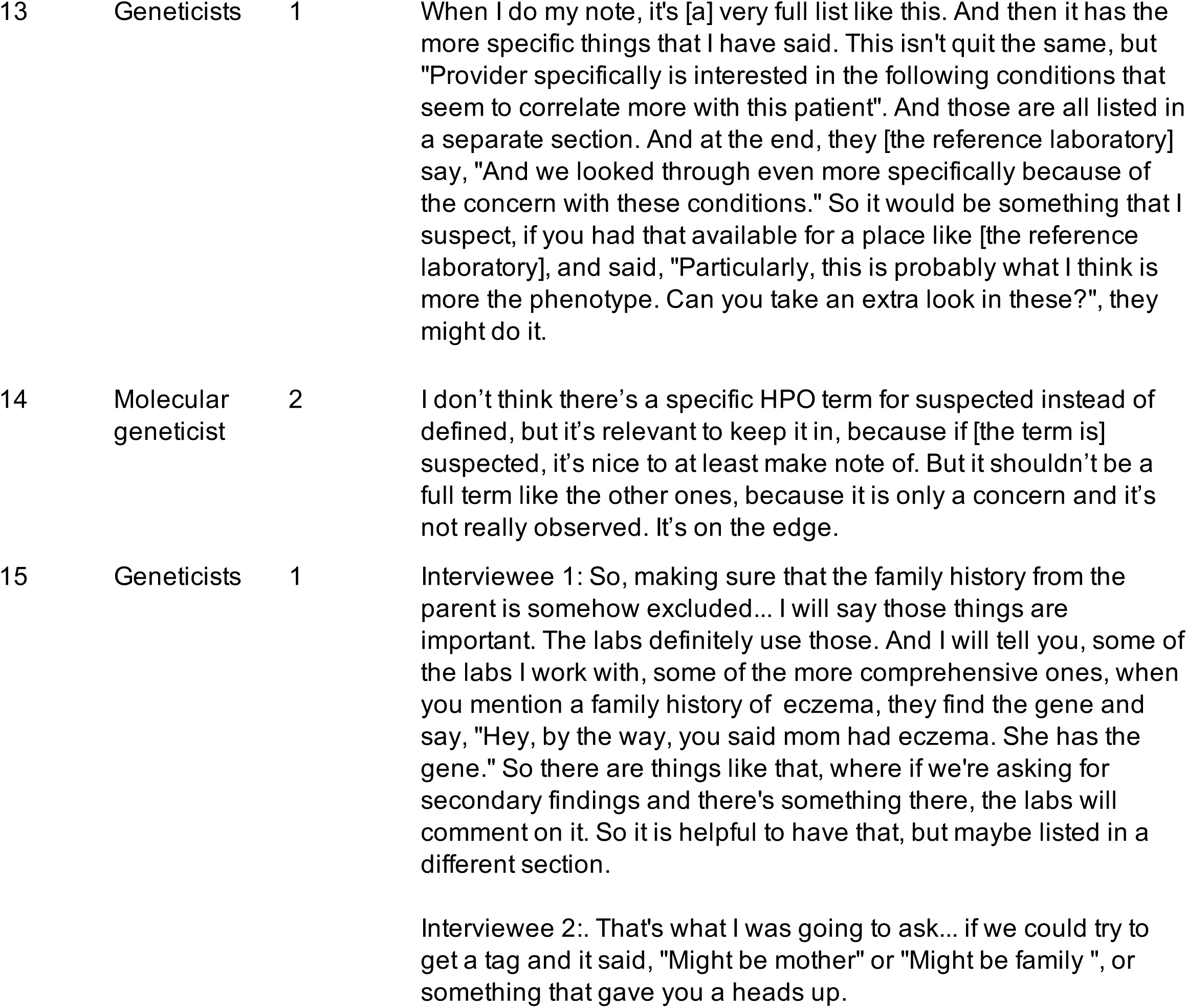
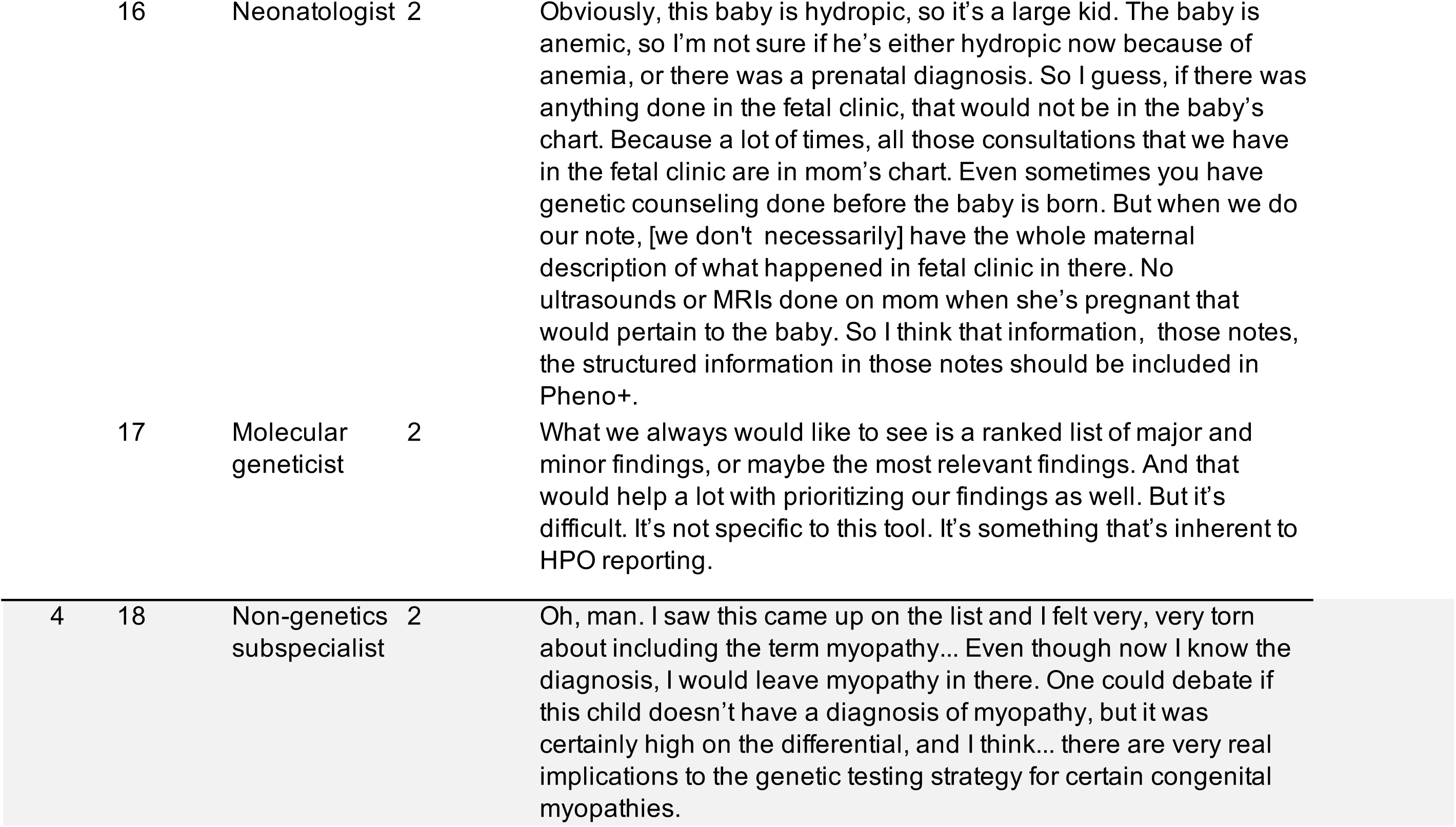
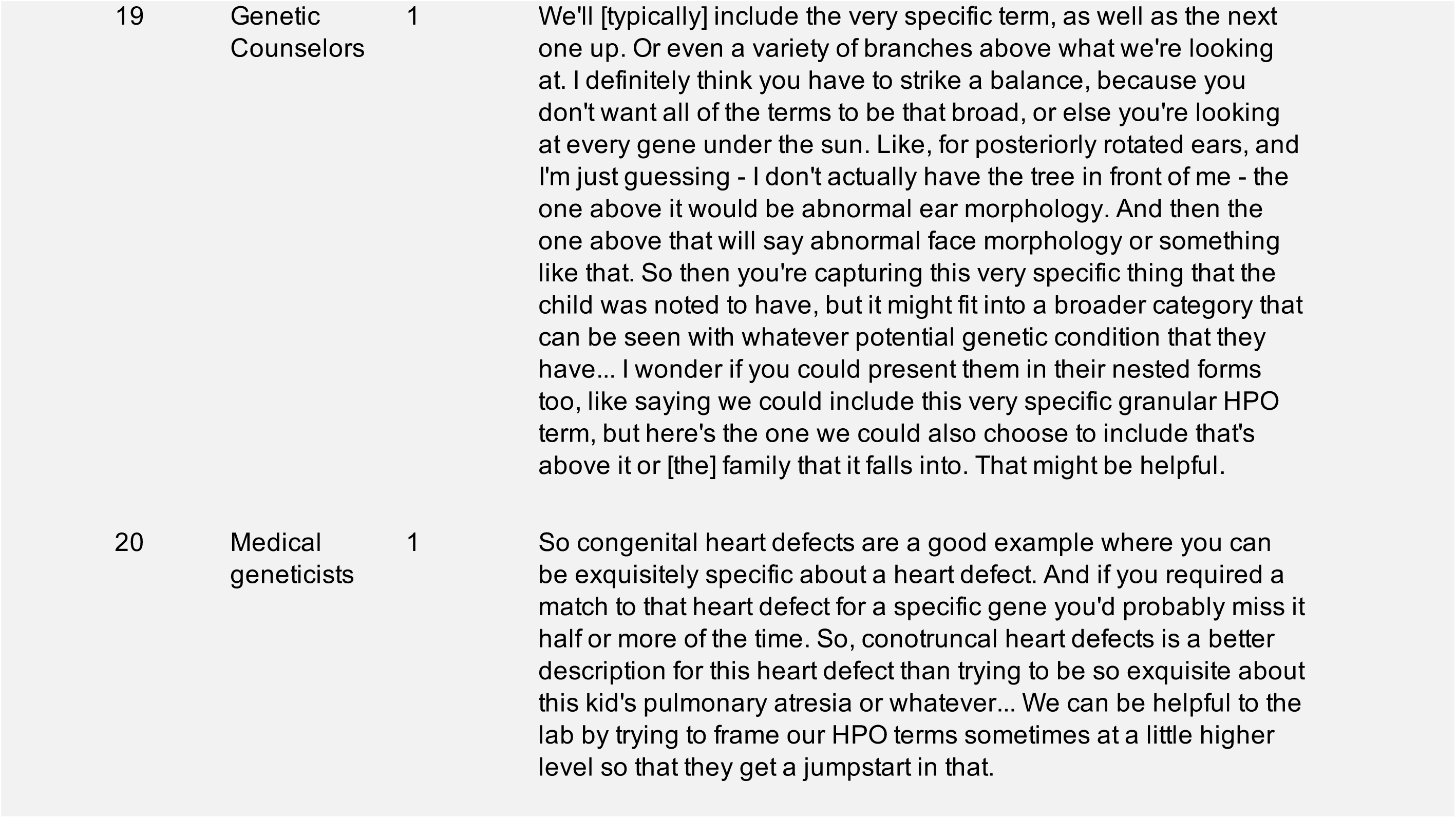
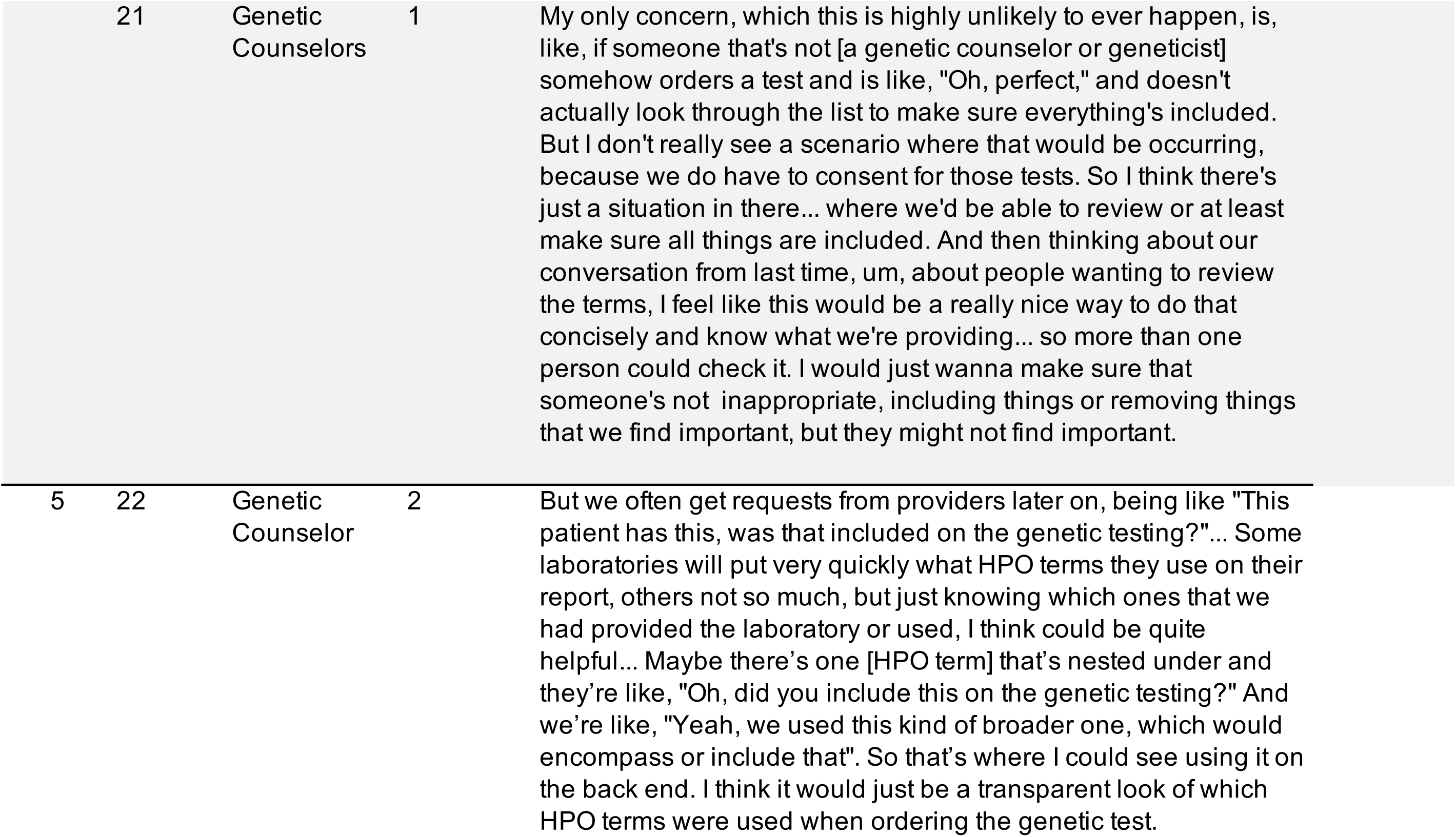
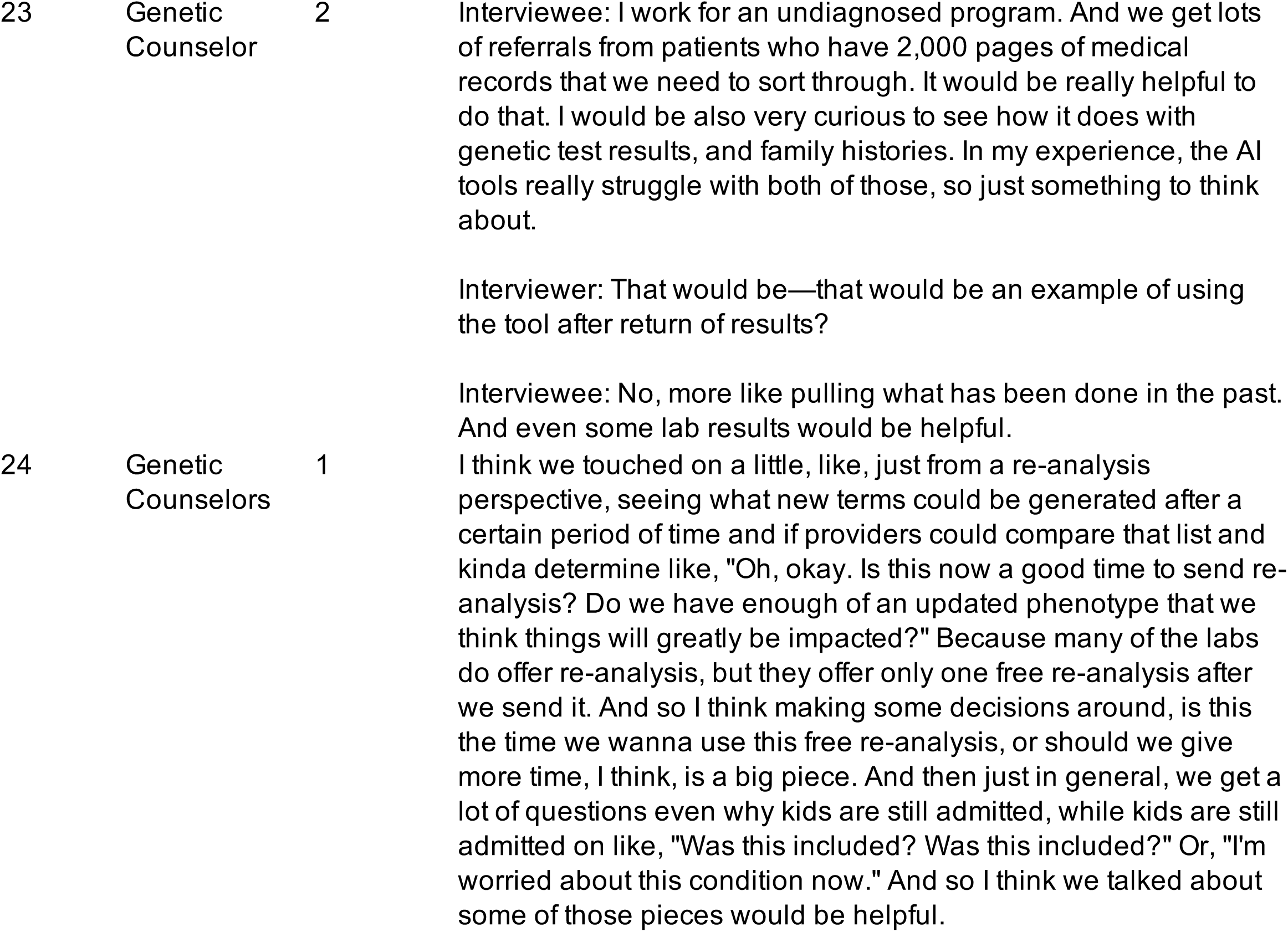
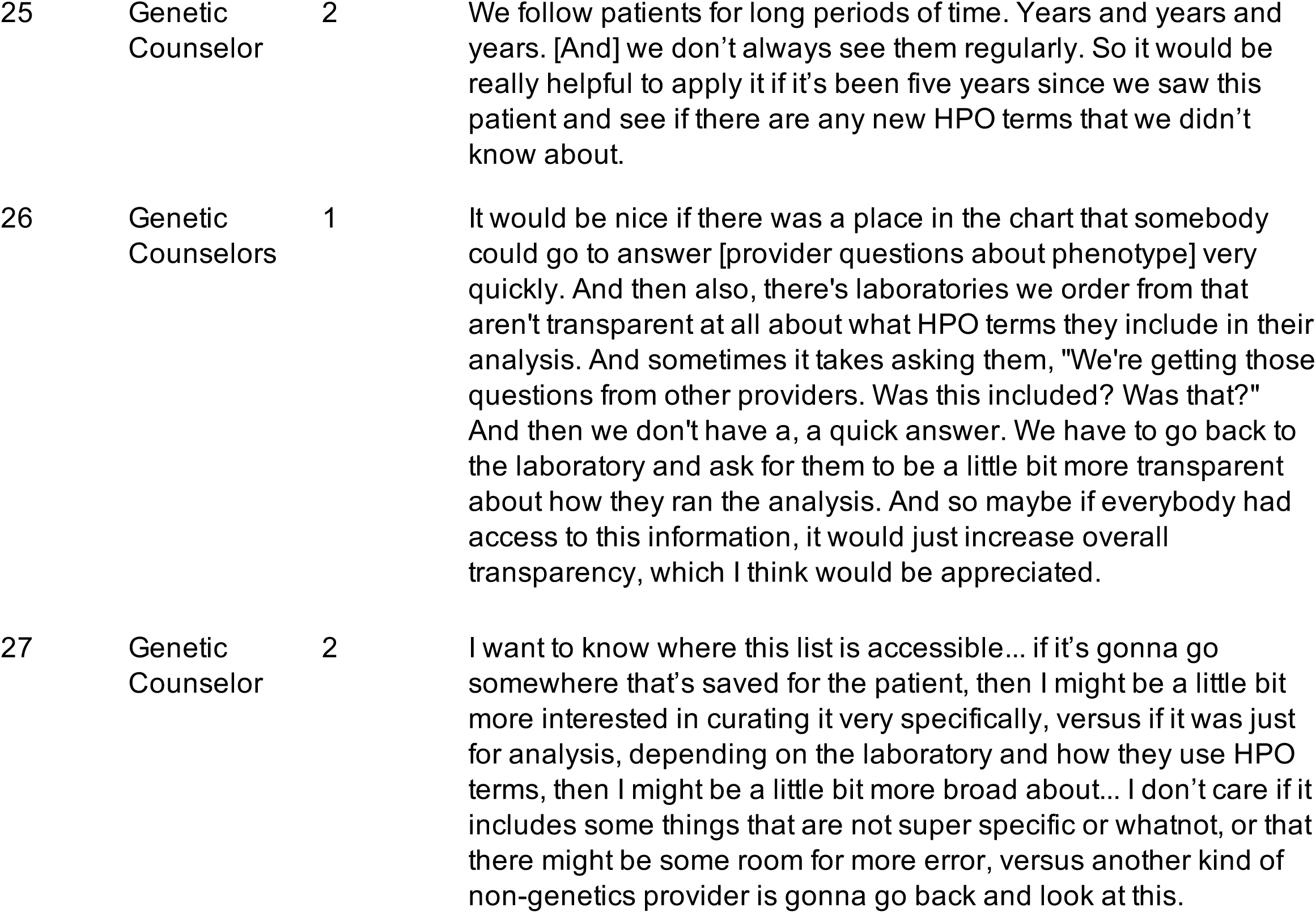

## Notes

### Competing Interest Statement

The authors have declared no competing interest.

### Author Declarations

In accordance with the Common Rule, the research was submitted to the University of Utah's Institutional Review Board and exempted (IRB_00165986).

## References

1. Hennekam RCM, Biesecker LG. Next-generation sequencing demands next-generation phenotyping. Human Mutation. 2012 2012;33(5):884–886. doi:10.1002/humu.22048

2. Baynam G, Walters M, Claes P, et al. Phenotyping: targeting genotype’s rich cousin for diagnosis. J Paediatr Child Health. Apr 2015;51(4):381–6. doi:10.1111/jpc.12705

3. Gargano MA, Matentzoglu N, Coleman B, et al. The Human Phenotype Ontology in 2024: phenotypes around the world. Nucleic Acids Research. 2024-01-05 2024;52(D1):D1333-D1346. doi:10.1093/nar/gkad1005

4. De La Vega FM, Chowdhury S, Moore B, et al. Artificial intelligence enables comprehensive genome interpretation and nomination of candidate diagnoses for rare genetic diseases. Genome Med. Oct 14 2021;13(1):153. doi:10.1186/s13073-021-00965-0

5. Shen F, Wang L, Liu H. Phenotypic Analysis of Clinical Narratives Using Human Phenotype Ontology. Studies in health technology and informatics. 2017 2017;245:581–585.

6. Maver A, Lovrecic L, Volk M, et al. Phenotype-driven gene target definition in clinical genome-wide sequencing data interpretation. Genet Med. Nov 2016;18(11):1102–1110. doi:10.1038/gim.2016.22

7. Lee JJY, van Karnebeek CDM, Wasserman WW. Development and user evaluation of a rare disease gene prioritization workflow based on cognitive ergonomics. J Am Med Inform Assoc. Feb 1 2019;26(2):124–133. doi:10.1093/jamia/ocy153

8. Slavotinek A, Prasad H, Yip T, Rego S, Hoban H, Kvale M. Predicting genes from phenotypes using human phenotype ontology (HPO) terms. Human Genetics. 2022-11-01 2022;141(11):1749-1760. doi:10.1007/s00439-022-02449-6

9. Johnson B, Ouyang K, Frank L, et al. Systematic use of phenotype evidence in clinical genetic testing reduces the frequency of variants of uncertain significance. Am J Med Genet A. Sep 2022;188(9):2642–2651. doi:10.1002/ajmg.a.62779

10. Kim S, Zhou Y, Guo Y, Xiao C, Zheng K. Applying natural language processing and large language models to clinical notes for phenotyping and diagnosing rare diseases: a systematic review. J Am Med Inform Assoc. Jun 1 2026;33(6):1225–1235. doi:10.1093/jamia/ocag045

11. Wiley K, Findley L, Goldrich M, et al. A research agenda to support the development and implementation of genomics-based clinical informatics tools and resources. J Am Med Inform Assoc. Jul 12 2022;29(8):1342–1349. doi:10.1093/jamia/ocac057

12. Bradshaw JM, Hoffman RR, Woods DD, Johnson M. The seven deadly myths of" autonomous systems". IEEE Intelligent Systems. 2013;28(3):54–61.

13. Dekker SW, Woods DD. MABA-MABA or abracadabra? Progress on human–automation co-ordination. Cognition, Technology & Work. 2002;4(4):240–244.

14. Hollnagel E. Flight decks and free flight: Where are the system boundaries? Decision making in aviation. Routledge; 2017:321–328.

15. Tsamados A, Floridi L, Taddeo M. Human control of AI systems: from supervision to teaming. AI and Ethics. 2025;5(2):1535–1548.

16. Woods D, Dekker S. Anticipating the effects of technological change: A new era of dynamics for human factors. Theoretical issues in ergonomics science. 2000;1(3):272–282.

17. Mandel JC, Kreda DA, Mandl KD, Kohane IS, Ramoni RB. SMART on FHIR: a standards-based, interoperable apps platform for electronic health records. J Am Med Inform Assoc. Sep 2016;23(5):899–908. doi:10.1093/jamia/ocv189

18. Vorisek CN, Lehne M, Klopfenstein SAI, et al. Fast Healthcare Interoperability Resources (FHIR) for Interoperability in Health Research: Systematic Review. JMIR Med Inform. Jul 19 2022;10(7):e35724. doi:10.2196/35724

19. 19. HL7. Resource DocumentReference. Accessed September 16, 2026. https://www.hl7.org/fhir/documentreference.html

20. Deisseroth CA, Birgmeier J, Bodle EE, et al. ClinPhen extracts and prioritizes patient phenotypes directly from medical records to expedite genetic disease diagnosis. Genet Med. Jul 2019;21(7):1585–1593. doi:10.1038/s41436-018-0381-1

21. Garcia BT, Westerfield L, Yelemali P, et al. Improving automated deep phenotyping through large language models using retrieval-augmented generation. Genome Med. Aug 18 2025;17(1):91. doi:10.1186/s13073-025-01521-w

22. Groza T, Gration D, Baynam G, Robinson PN. FastHPOCR: pragmatic, fast, and accurate concept recognition using the human phenotype ontology. Bioinformatics. Jul 1 2024;40(7)doi:10.1093/bioinformatics/btae406

23. Luo L, Yan S, Lai PT, et al. PhenoTagger: a hybrid method for phenotype concept recognition using human phenotype ontology. Bioinformatics. Jul 27 2021;37(13):1884–1890. doi:10.1093/bioinformatics/btab019

24. Yang J, Liu C, Deng W, et al. Enhancing phenotype recognition in clinical notes using large language models: PhenoBCBERT and PhenoGPT. Patterns (N Y). Jan 12 2024;5(1):100887. doi:10.1016/j.patter.2023.100887

25. Brooke J. SUS: a retrospective. Journal of usability studies. 2013;8(2):29.

26. Braun V, Clarke V. Using thematic analysis in psychology. Qualitative research in psychology. 2006;3(2):77–101.

27. Harvey G, Kitson A. PARIHS revisited: from heuristic to integrated framework for the successful implementation of knowledge into practice. Implement Sci. Mar 10 2016;11:33. doi:10.1186/s13012-016-0398-2

28. Deisseroth CA, Birgmeier J, Bodle EE, et al. ClinPhen extracts and prioritizes patient phenotypes directly from medical records to expedite genetic disease diagnosis. Genetics in Medicine. 2019 2019;21(7):1585–1593.

29. Bonet A, Tort-Nasarre G, Domènech-Sorolla J, Monistrol O, Camí C, Medel D. "Perceptions of artificial intelligence in nursing students: A qualitative meta-synthesis based on the UTAUT2 model". Nurse Educ Today. Aug 2026;163:107119. doi:10.1016/j.nedt.2026.107119

30. Jørgensen NL, Bruun NH, Hoffmann Merrild C, Moeslund TB, Kidholm K, Thomsen JL. Identifying factors influencing acceptance of artificial intelligence among general practitioners in Danish general practice: a cross-sectional web-based survey study. Scand J Prim Health Care. Dec 2026;44(1):2667990. doi:10.1080/02813432.2026.2667990

31. Gallant NL, Hadjistavropoulos T, Stopyn RJN, Feere EK. Integrating Technology Adoption Models Into Implementation Science Methodologies: A Mixed-Methods Preimplementation Study. Gerontologist. Mar 21 2023;63(3):416–427. doi:10.1093/geront/gnac098

32. Venkatesh V, Morris MG, Davis GB, Davis FD. User acceptance of information technology: Toward a unified view1. MIS quarterly. 2003;27(3):425–478.

33. Ladewig MS, Jacobsen JOB, Wagner AH, et al. GA4GH Phenopackets: A Practical Introduction. Adv Genet (Hoboken). Mar 2023;4(1):2200016. doi:10.1002/ggn2.202200016

34. Wilson E, Daniel M, Rao A, et al. A scoping review of distributed cognition in acute care clinical decision-making. Diagnosis (Berl). May 1 2023;10(2):68–88. doi:10.1515/dx-2022-0095

35. Hutchins E. How a cockpit remembers its speeds. Cognitive science. 1995;19(3):265–288.

36. Goodwin C. Co-operative action. Cambridge University Press; 2018.

37. Griffen Z, Asfaha DM, Owens K. The Biggest Struggle: Navigating Trust and Uncertainty in Genetic Variant Interpretation. Public health genomics. 2024;27(1):228–232.

38. Ackerman SL, Koenig BA. Understanding variations in secondary findings reporting practices across U.S. genome sequencing laboratories. AJOB Empir Bioeth. Jan-Mar 2018;9(1):48–57. doi:10.1080/23294515.2017.1405095

39. Balcarcel DR, Mehta SD, Dixon CG, et al. Feedback loops in intensive care unit prognostic models: an under-recognised threat to clinical validity. Lancet Digit Health. Aug 2025;7(8):100880. doi:10.1016/j.landig.2025.100880

40. Butler JM, Taft T, Taber P, et al. Pneumonia diagnosis performance in the emergency department: a mixed-methods study about clinicians’ experiences and exploration of individual differences and response to diagnostic performance feedback. J Am Med Inform Assoc. Jun 20 2024;31(7):1503–1513. doi:10.1093/jamia/ocae112

41. Croskerry P. The feedback sanction. Academic Emergency Medicine. 2000;7(11):1232–1238.

42. Greenhalgh T, Abimbola S. The NASSS framework-a synthesis of multiple theories of technology implementation. Stud Health Technol Inform. 2019;263(263):193–204.

43. Greenhalgh T, Wherton J, Papoutsi C, et al. Beyond adoption: a new framework for theorizing and evaluating nonadoption, abandonment, and challenges to the scale-up, spread, and sustainability of health and care technologies. Journal of medical Internet research. 2017;19(11):e8775.

44. Skraaning Jr G, Jamieson GA. The failure to grasp automation failure. Journal of Cognitive Engineering and Decision Making. 2024;18(4):274–285.

45. Bainbridge L. Ironies of automation. Analysis, design and evaluation of man– machine systems. Elsevier; 1983:129–135.

