## Supplementary material for "Usability and workflow implications of an electronic health record-integrated tool for automated phenotyping: Formative evaluation of Pheno+": Interview guide

***Pheno+ evaluation Round 2 guide***

Thank you for agreeing to do this user-centered design session with us. We will ask you to use and provide some feedback on a new tool, called Pheno+ tool. ***Pheno+*** is a tool for generating and curating Human Phenotype Ontology (HPO) terms from a patient’s clinical notes directly within the EHR. The tool is designed to allow providers or clinical staff to independently generate a list of phenotypes that describe a patient’s presentation using the EHR notes already generated by the interdisciplinary clinical team. Phenotype terms that are generated in Pheno+ can then be copied and used as an input for downstream case analysis tools.

In this session, we will ask you to use the tool to generate phenotypes for 3 patients, and to explain a bit about your reasoning as you do so. We expect that this interview will take about 45 minutes. We will digitally record the session so that we can understand your responses as clearly as possible.

You should have received a DocuSign consent document. If you haven’t already, please take a moment to find that document in your email, review and sign it.

Do you have any questions?

*[Begin recording]*

*Accessing the Application*

1. Please log into Epic.
2. Go to Patient Lookup and enter MRN [number]
3. Go to Hospital Admission.
4. Once within the patient chart use the upper “search” bar in the Epic header and begin typing “pheno+”. As you type “Pheno+” should appear as an option below under Patient Activities in the dropdown. Select it by clicking it to open the app.
5. This will load your patient case’s notes within the app

*Using the Application*

- You should see the available notes for this case as a starting point within the Pheno+ app. This is located on the left-hand side in the “relevant EHR notes” section.
- Directly to the right of that section is a collapsable preview of the currently selected note’s content. Closing this window will allow more space for the relevant EHR notes section and will allow you to see the full specialty and other information about the note.
- Many sections of this app allow you to collapse or expand them for more screen real estate as needed. Generally, this can be achieved by clicking a button near the top of that section.
- Under relevant EHR notes, you can choose which notes you want to include. And process them into HPO terms. As you do this the term review section will open to allow you to curate your list down to relevant terms.
- There are additional term interrogation capabilities within the term review section. Namely you can expand and review the note context that the term was generated from to assess its relevance. You can also see the approximate number of times that term was seen.
- Please add or remove terms as necessary until you reach a satisfactory list of HPO terms.
- I will ask you some questions about your actions as you proceed to construct a list of HPO terms.

*[Questions after EACH PATIENT]*

- *[IF UNADDRESSED DURING THINK-ALOUD]* Did you see any terms indicated by Pheno+ that appear to be errors?
  - [if YES] How did you recognize errors?
- *[IF UNADDRESSED DURING THINK-ALOUD]* Were there any terms you would have liked to add but couldn’t find?

Next, I have a few statements about how much you trust the Pheno+ tool. For each statement, we’d like you to give a rating between 1 and 5, where 1 is “strongly disagree”, 3 is “neutral” and 5 is “strongly agree”.

- I trust the tool output in this case.
- I would ~~have~~ manage this case better without the tool output.
- In this case, the tool performed well enough to save me time.
- Relying on the tool output in this case would be risky.
- Ignoring the tool output could lead me to an unsafe judgment in this case.
- [IF UNADDRESSED] What are the main factors influencing your trust in the tool?

*[Questions after COMPLETING ALL PATIENTS]*

- Please share what your experience has been like using Pheno+ overall.
- What did you like most about interacting with the tool?
- What did you like least?
- Did you notice any differences in the usefulness of the tool for the different patients you looked at?
- What questions do you have about how the tool works?
  - Do these questions impact how you would use the tool?
- At what points in your workflow do you envision possibly using Pheno+?
  - Are there portions of the workflow after return of results when you think Pheno+ might be useful?

*[Questions for DIAGNOSTIC SPECIALIST/[MOLECULAR] GENTICIST]*

- Would you find the output of this tool useful if it was sent to your laboratory with the sample and test requisition form?
  - How would you use it?

Thank you for reviewing the Pheno+ tool! To complete this evaluation, we ask you to complete a survey about your experience, which you can find at the link in our chat, or in an email you should have received from [NAME].

[SURVEY LINK]

Upon completion, we will be providing a $50 gift card for you as a token of our appreciation for your participation in this study. Thank you!
